# Early detection of mandible osteoradionecrosis risk in a high comorbidity Veteran population

**DOI:** 10.1101/2022.07.01.22277008

**Authors:** David Chamberlayne Wilde, Sagar Kansara, Lucas Gomez Sapienza, Logan Banner, Rickey Morlen, David Hernandez, Andrew Tsao Huang, Weiyuan Mai, Clifton David Fuller, Stephen Lai, Vlad Constantin Sandulache

## Abstract

**Objective:** Osteoradionecrosis (ORN) of the mandible is a devastating complication of external beam radiation therapy (EBRT) for head and neck squamous cell carcinoma (HNSCC). We sought to ascertain ORN risk in a Veteran HNSCC population treatment with definitive or adjuvant EBRT and followed prospectively.

**Study design:** Retrospective analysis of prospective cohort.

**Setting:** Tertiary care Veterans Health Administration (VHA) medical center.

**Methods:** Patients with HNSCC who initiated treatment at the Michael E. DeBakey Veterans Affairs Medical Center (MEDVAMC) are prospectively tracked for quality of care purposes through the end of the cancer surveillance period (5 years post treatment completion). We retrospectively analyzed this patient cohort and extracted clinical and pathologic data for 164 patients with SCC of the oral cavity, oropharynx, larynx and hypopharynx who received definitive or adjuvant EBRT (2016-2020).

**Results:** Most patients were dentate and 80% underwent dental extractions prior to EBRT of which 16 (16%) had complications. The rate of ORN was 3.7% for oral cavity SCC patients and 8.1% for oropharyngeal SCC patients. Median time to ORN development was 156 days and the earliest case was detected at 127 days post EBRT completion. All ORN patients were dentate and underwent extraction prior to EBRT start.

**Conclusion:** ORN development can occur early following EBRT in a Veteran population with significant comorbid conditions but overall rates are in line with the general population. Prospective tracking of HNSCC patients throughout the post-treatment surveillance period is critical to early detection of this devastating EBRT complication.

## Introduction

Osteoradionecrosis (ORN) of the craniofacial skeleton is a feared complication of external beam radiation therapy (EBRT) that can lead to devastating health effects for patients treated for a diagnosis of head and neck squamous cell carcinoma (HNSCC).^1^ Precise definitions vary, but all describe an area of chronically exposed bone in the irradiated field that does not heal in the absence of recurrent neoplastic disease.^2^ The pathophysiology of ORN was once believed to stem from a triad of radiation, trauma, and infection.^3^ Reports of the development of disease in the absence of one or more of these factors prompted investigation of other explanations.^4^ Revised theories on the etiology of ORN take into consideration a multifactorial generation of tissue hypoxia stemming from small vessel compromise and hypocellularity along with induction of fibrosis by reactive oxygen species as a late complication of high dose radiation leading to replacement of parenchymal tissue with dense extracellular matrix. Imaging studies conducted by the Joint Head and Neck Radiotherapy-MRI Development Cooperative at the University of Texas MD Anderson Cancer Center and collaborating institutions strongly support a hypothesis of radiation-altered bone vascularity as a critical contributor to ORN development.^**5-7**^

Reported ORN rates range from 0-16%.^8-10^ This discrepancy in disease incidence may be partially explained by the evolution of radiation delivery from three dimensional conformal strategies to intensity-modulated radiation therapy (IMRT), which can deliver comparable doses to tumors with lower radiation delivery to surrounding tissues.^11^ This assumption, though widely described, is challenged by several retrospective analyses.^10,12^ Although more conformal radiation planning would be expected to provide improved bone sparing, it is unclear whether or not *any* dose conformation can spare the ipsilateral body and angle of the mandible for oropharyngeal tumors with concomitant nodal disease.^13^ It is therefore unlikely that technical improvements in dose delivery alone can eliminate ORN risk, particularly in high risk patient populations with a high comorbidity burden that could impact soft tissue and bony healing.^14,15^.

For the current analysis we leveraged a tracking tool of treatment delivery for high-comorbidity (complicated diabetes mellitus, hypertension, peripheral vascular disease)^16,17^, high-risk Veterans with a diagnosis of HNSCC as previously reported by our group.^18^ We sought to characterize the incidence of ORN at a tertiary Veterans Health Administration (VHA) institution with on-site Dental and Oral and Maxillofacial Surgery services integrated into a multi-disciplinary treatment planning group and identify correlates of ORN development.

## Materials and methods

We retrospectively analyzed prospectively collected data from an institutional cohort of patients treated with curative intent EBRT for HNSCC in the definitive or adjuvant setting. Following approval from Baylor College of Medicine and Michael E. DeBakey Veterans Affairs Medical Center (MEDVAMC) Institutional Review Boards, we reviewed the records of patients who were diagnosed and initiated radiation therapy at the MEDVAMC between January 2016 and December 2020 captured in our treatment tracking registry.^18^ Patient demographics, dental records and procedures, tumor and treatment characteristics, and data pertaining to the development and treatment of ORN were retrospectively obtained from the existing electronic medical record and the registry. T, N, M classification were recorded according to the American Joint Commission on Cancer (AJCC Staging Manual 8^th^ Edition) staging system. Associations between clinical variables and the development of ORN were investigated using two-directional Fisher’s exact tests for nominal and Mann-Whitney U for ordinal data. Charlson Comorbidity Index (CCI) was calculated using criteria published by Charlson *et al*.^19^

## Results

### Patient characteristics

Our study encompassed 164 HNSCC patients with an average age of 65 years (**Table 1**); 158 (96.3%) were male. The median follow-up time was 725 days (range 49-1817 days). Most patients (142; 87%) were current or former smokers, with a mean pack year history of 47.2; 119 had a history of alcohol consumption (73%). The average CCI was 5.47 and 40 (24%) patients had a documented history of diabetes mellitus. Only 122 (74.4%) patients were dentate.

**Table 1.** Patient characteristics

| **Table 1. Patient characteristics** |  |  |  |  |
| --- | --- | --- | --- | --- |
|  |  | **All patients** | **non-ORN** | **ORN** |
| Age | Mean/SD (years) | 65.3 / 7.82 | 65.3 / 7.72 | 64.9 / 10.5 |
| CCI | Mean/SD | 5.47 / 1.99 | 5.51 / 2.01 | 4.57 / 1.13 |
| Tobacco exposure |  | N (%) | N (%) | N (%) |
|  | former | 142 (86.6) | 137 (87.3) | 5 (71.4) |
|  | current | 69 (42.1) | 67 (42.7) | 2 (28.6) |
| Alcohol consumption | former | 119 (72.6) | 113 (72.0) | 6 (85.7) |
|  | current | 62 (37.8) | 59 (37.6) | 3 (42.9) |
| HPV/p16 status | positive | 66 (40.2) | 61 (38.9) | 5 (71.4) |
|  | negative | 98(59.8) | 96 (61.1) | 2 (28.6) |
| Diabetes mellitus |  | 40 (24.4) | 37 (23.6) | 3 (42.9) |
| Dentate |  | 122 (74.4) | 115 (73.2) | 7 (100) |

Note- ORN: osteoradionecrosis. SD: standard deviation. HPV: human papillomavirus. CCI: Charlson Comorbidity Index.

### Tumor and treatment characteristics

Primary tumor site distribution was as follows: 27 (16.4%) oral cavity, 78 (47.5%) oropharynx, 6 (3.7%) hypopharynx, and 53 (32.3%) larynx. The majority of patients (79, 52.6%) presented with T1-T2 tumors, largely driven by a high frequency of early T-classification laryngeal disease (**Table 2**). Concomitantly, most (113, 68.9%) patients had N0 or N1 nodal disease at the time of diagnosis. All patients underwent primary or adjuvant treatment with EBRT as recommended by the institutional multidisciplinary tumor board and relevant National Comprehensive Cancer Network (NCCN) guidelines; all patients were treated with IMRT with a mean dose of 65.4Gy. While almost all (96.3%) oral cavity tumors underwent surgical resection, 70 (89.7%) oropharyngeal tumors were treated with primary EBRT with or without chemotherapy depending on extent of disease (**Table 3**).

**Table 2.** Tumor Characteristics and Treatment by Site

| Table 2. Tumor Characteristics and Treatment by Site |  |  |  |  |  |  |  |
| --- | --- | --- | --- | --- | --- | --- | --- |
|  | T1-T2<br>N (%) | T3-T4<br>N (%) | N0-N1<br>N (%) | >N1<br>N (%) | Primary<br>Surgery N (%) | Primary EBRT<br>N (%) | Chemotherapy<br>N (%) |
| All sites | 79 (48.2) | 85 (51.8) | 113 (68.9) | 51 (31.1) | 47 (28.7) | 117 (71.3) | 101 (61.6) |
| OC | 10 (37.0) | 17 (63.0) | 16 (59.3) | 11 (40.7) | 26 (96.3) | 1 (3.70) | 13 (48.1) |
| OP | 31 (39.7) | 47 (60.3) | 48 (61.5) | 30 (38.5) | 8 (10.3) | 70 (89.7) | 72 (92.3) |
| HP | 2 (33.3) | 4 (66.7) | 3 (50) | 3 (50) | 1 (16.7) | 5 (83.3) | 5 (83.3) |
| LA | 36 (67.9) | 17 (32.1) | 46 (86.8) | 7 (13.2) | 12 (22.6) | 41 (77.4) | 11 (20.8) |

Note- OC = oral cavity; OP = oropharynx; HP = hypopharynx; LA = nasopharynx

**Table 3.** Extraction Data

| Table 3. Extraction Data |  |  |  |  |
| --- | --- | --- | --- | --- |
|  |  | All patients (%) | Non-ORN (%) | ORN (%) |
| Dentate (pre-EBRT) |  | 122 (74.4) | 115 (73.2) | 7 (100) |
| Pre-EBRT extraction |  | 98 (59.8) | **91 (58.0)** | **7 (100)** |
| Extraction complications |  | 16 (9.76) | 13 (8.28) | 3 (42.6) |
| Teeth Extracted | Mean/SD | 13.7 / 8.96 | 13.7 / 9.08 | 13.7 / 7.93 |
| Extraction to EBRT | Mean/SD (days) | 30.4 / 11.5 | 30.7 / 11.6 | 26.6 / 9.29 |

Note- EBRT: external-beam radiation therapy. ORN: osteoradionecrosis. Bold italicized numbers indicated significant differences between ORN and non-ORN cohorts ( $p < 0.05$ , Z-score).

### Dental status pre-EBRT

Of the 122 patients that were dentate prior to initiation of EBRT, 98 (80.3%) underwent prophylactic dental extractions; 74 of these consisted of removal of all remaining teeth. The mean number of teeth extracted was 13.7. Sixteen patients had documented post-extraction wound complications ranging from food impaction to exposed bone. The mean time from dental extraction to the initiation of EBRT was 30.4 days (range 5-83 days).

### ORN development

The overall rate of development of ORN in our cohort was 4.27%. Of the 7 patients who developed ORN (**Table 4**), one had an oral cavity primary with the rest originating in the oropharynx and one p16+ SCC defined as a carcinoma of unknown primary (CUP). The rate of ORN development by subsite was 3.7% for oral cavity, 8.1% for oropharynx, and 0% for hypopharyngeal and laryngeal tumors. The mean time from the start of EBRT to development of ORN was 196 days with a median of 156 days (range 127-368 days). All 7 patients had involvement of the mandible and all patients with known primary tumor laterality had involvement of the ipsilateral mandible (**Figure 1**). Six patients had radiographic evidence of ORN at the time of clinical appearance; four patients demonstrated significant bony erosion (**Figure 2**) Five were smokers with an average of 47.2 pack years; 1 patient continued to smoke during EBRT. Three had documented complications with pre-radiation dental extractions with exposed bone that slowly re-mucosalized prior to initiation of treatment.

**Table 4.** ORN cases

| Table 4. ORN cases |  |  |  |  |  |  |  |  |  |  |
| --- | --- | --- | --- | --- | --- | --- | --- | --- | --- | --- |
| Patient | Site | Subsite Stage | ORN | p16 | Pack-years | Extractions | Dental complications | EBRT dose primary/neck (Gy) | EBRT – ORN (days) | Treatment |
| 1 | OC | buccal T2N0M0 | mandibular ramus (ipsilateral) | - | 55 | y |  | 60/60 | 148 | HBO, surgery |
| 2 | OP | tonsil T4N3M0 | mandibular body (ipsilateral) | + | 10 | y | y | 70 | 368 | HBO, surgery |
| 3 | OP | Tonsil T3N3bM0 | mandibular body (ipsilateral) | - | 100 | y |  | 66/59 | 156 | surgery |
| 4 | OP | Tonsil T1N1M0 | mandibular angle (ipsilateral) | + | 11 | y | y | 70/63 | 205 | surgery |
| 5 | OP | tonsil T4N1M0 | mandibular body (ipsilateral) | + | 0 | y | y | 70 | 127 | non-surgical |
| 6 | OP | tongue base T4N1M0 | mandibular body (ipsilateral) | + | 0 | y |  | 70/63 | 224 | surgery |
| 7 | OP | CUP | mandibular angle (NA) | + | 60 | y |  | 70/63 | 147 | surgery |

**Figure 1.**
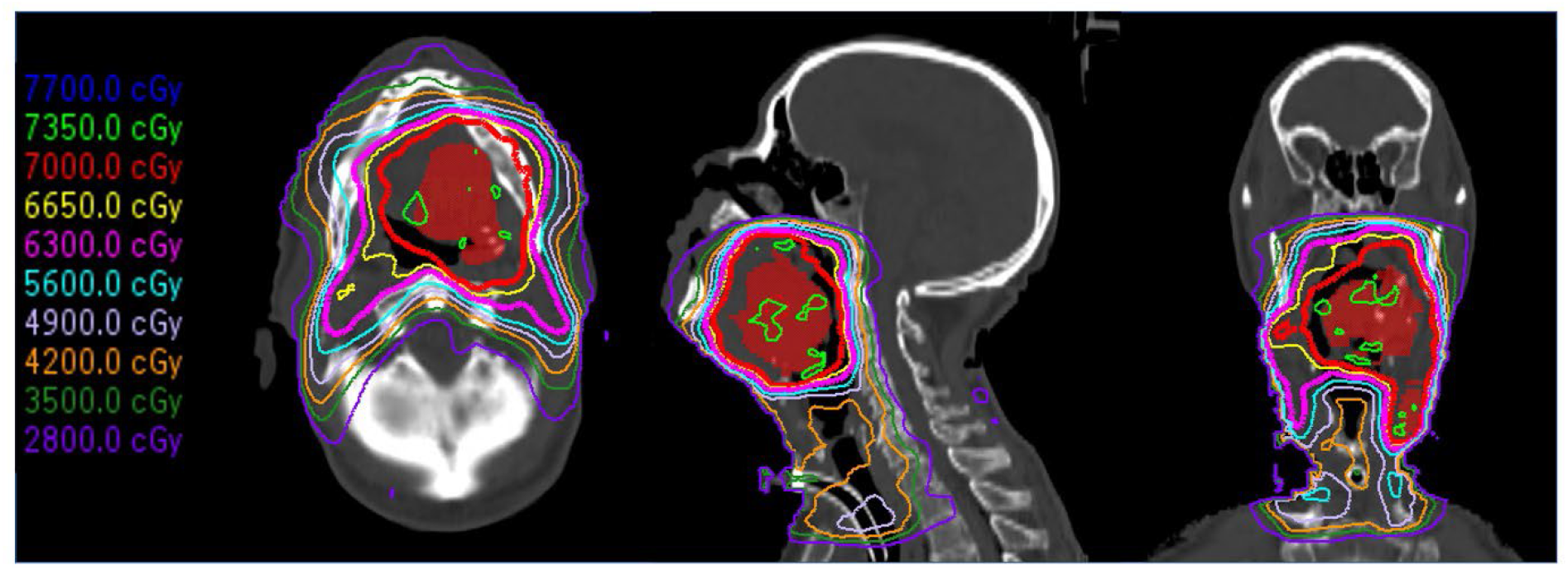
Radiation plan for oropharyngeal cancer. Dosimetric plan for T4N2M0 OPSCC which developed right mandibular ORN. Approximately 75% of the mandibular volume falls within 56Gy dose and approximately 90% falls within 35Gy dose.

**Figure 2.**
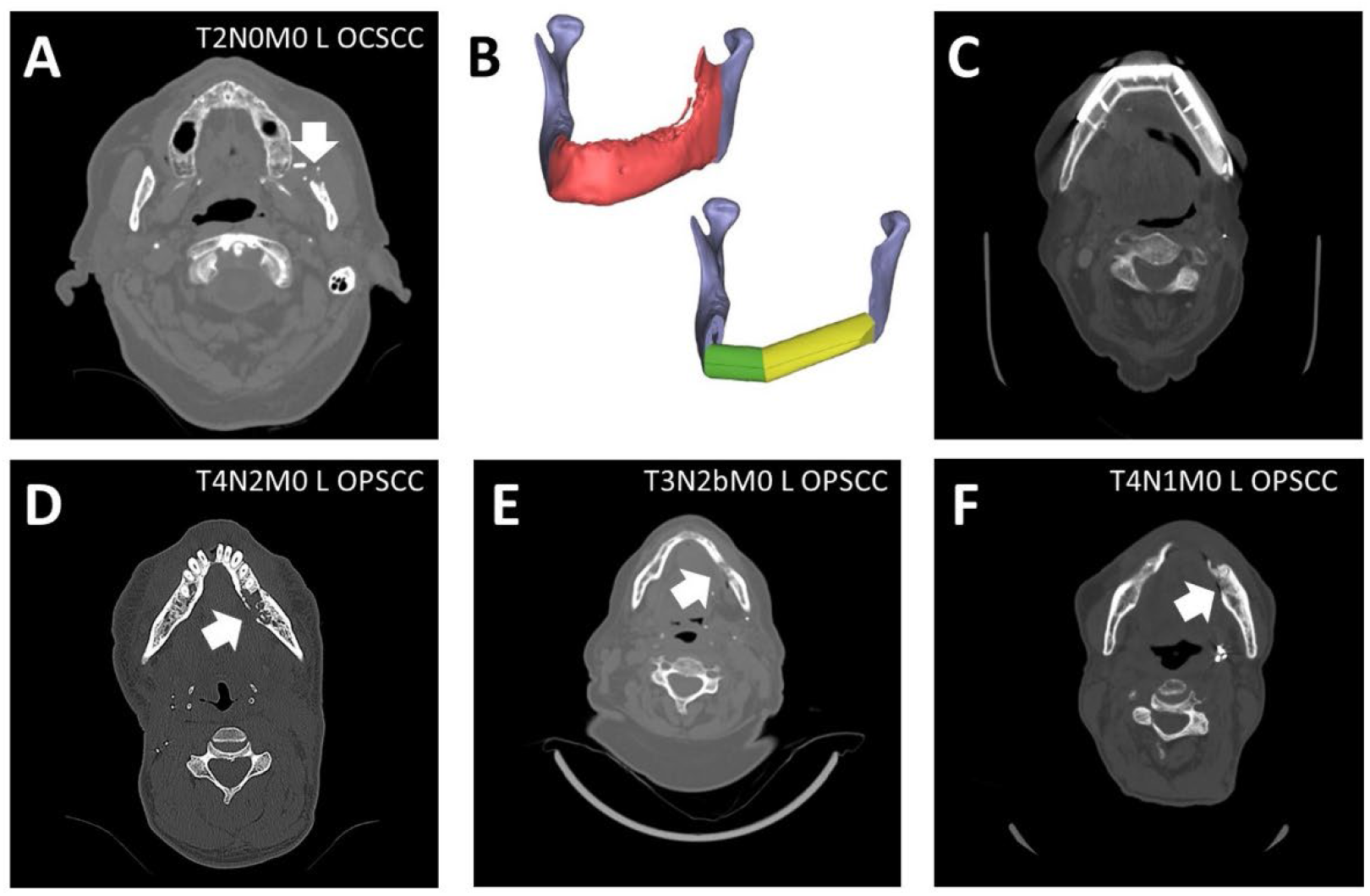
ORN development. A) Erosion of the mandibular body, reconstructed using a fibular free flap. B) Operative planning for reconstruction. C) Imaging of reconstructed mandible. D-F) Bony erosion at the time of diagnosis.

### ORN treatment

All patients were treated with aggressive oral hygiene and were evaluated by the Otolaryngology-Head and Neck Surgery (Oto-HNS) and Oral Maxillofacial Surgery (OMFS) teams. Two patients underwent hyperbaric oxygen (HBO), and a third was offered this therapy, but had a distant recurrence at the time of diagnosis and opted to forgo it. Three patients underwent segmental mandibulectomy reconstructed as follows: fibula free flap (n=1), soft tissue free flap (n=1), pectoralis flap (n=1) (aborted fibula free flap due to inadequate recipient vessels). A fourth patient underwent operative debridement and reconstruction of the mucosal defect with a facial artery myomucosal flap (**Figure 2**). There were no free flap failures. The patient reconstructed with a pectoralis flap had a post-operative course complicated by orocutanous fistula which healed with local wound care.

## Discussion

Over the last three decades, a new epidemic of human papilloma virus (HPV) driven oropharyngeal squamous cell carcinoma (SCC) has changed the landscape of patients receiving head and neck EBRT.^20^ This has led to both increasing numbers of patients receiving EBRT at time of diagnosis, as well as more patients surviving long enough to be effected by late complications of this therapy.^21,22^ It is now clear that HPV+OPSCC patients are generally younger and healthier compared to their HPV-counterparts.^23^ When superimposed on a substantially lower tobacco exposure, it is likely that HPV+OPSCC patients may constitute a “low-risk” cohort for EBRT related complications, particularly development of ORN.^21^ In contrast, Veteran patients present not only with a disproportionately higher incidence of HPV-disease including OPSCC but a disproportionately higher comorbidity burden.^16,17,24-28^ This is particularly relevant to ORN development which may be impacted by comorbidities for which Veterans are particularly enriched such as peripheral vascular disease and diabetes mellitus.^26,28^

The overall rate of ORN in our cohort is within the range of reported incidence in the literature. As shown here, no patients with laryngeal SCC developed ORN, as would be expected with a fairly high complement of early-stage disease resulting in narrower radiation fields and complete sparing of the mandible. This finding is important because it has allowed us to tailor both our pre-treatment patient counseling and our post-EBRT bone surveillance algorithms accordingly. In contrast, 8% of patients with OPSCC developed ORN. Despite the promise of IMRT and even intensity-modulated proton beam therapy (IMPT), it is not technically possible to protect the mandible when delivering curative doses of radiation to the primary tumor and nodal areas.^29^ Comparisons among patient cohorts therefore must account for primary disease site, subsite, and T- and N-classification as ORN rates are likely to be very significantly impacted by these parameters. In contrast to our cohort, many recent datasets are skewed towards less locally advanced disease.^30-32^ A recent study from the University of Texas MD Anderson Cancer Center described an ORN rate of 13.7% among all HNSCC patients with a 5% rate of grade 4 ORN. A subset analysis generated for the current manuscript demonstrated that 123/826 oropharyngeal cancer patients (14.9%) developed all grade ORN and 42/825 (5.1%) developed grade 4 ORN.^33^ In the current cohort, Veterans who developed ORN were more likely to have pre-EBRT extractions (Table 3). No other variables reached statistical significance (Tables 1, 3).

Given the high burden of comorbidities relevant to mandibular vascularity and tobacco exposure in our Veteran cohort, it is somewhat re-assuring to uncover an ORN rate within the reported literature range. A notable finding in our cohort is the short interval between the delivery of EBRT and the development of ORN (median time to ORN = 156 days). This is substantially more rapid that intervals reported in other studies.^10^ Although our median time to ORN is undoubtedly impacted by our short median follow up time as is the rate potentially, it is important to note that time to earliest ORN (127 days) is unlikely to change and strongly suggests the need for careful and proactive tracking of patients throughout both treatment delivery and the surveillance period following treatment completion.

A possible explanation for early ORN development is the underlying poor dental health of our patients. As previously stated, one of the factors that puts patients at highest risk for the development of ORN is the breakdown of barrier mechanisms from infection by the extraction of teeth in the irradiated field. All ORN patients were dentate pre-EBRT, underwent extractions and 3/7 developed post extraction complications. Perhaps either the poor pre-existing state of the mucosal and/or alveolar structures of the jaw or the maladaptive oral hygiene habits of our patients initiated a more rapid evolution of disease. A decreased time to insult of these local tissues could have started an earlier cascade of demucosalization and infection leading to bone death. Alternatively, since our HNSCC patients are tracked prospectively pre- and post-treatment, this time to development could in fact be more accurate compared to other purely retrospective analyses. All of our dentate HNSCC patients at the MEDVAMC undergo evaluation by the Dental and OMFS teams for optimization of oral health and possible extractions prior to initiation of radiation therapy. Although very few patients develop complications from extractions, these complications can result in treatment initiation delays and correlate with ORN risk. Nevertheless, because patients are tracked prospectively in real time, continued communication between the treatment teams can still minimize treatment delays and interruptions.

There are several limitations to the currently study, chiefly the sample size of patients enrolled. Although the MEDVAMC contains the highest volume Otolaryngology-Head and Neck Service Line within the VHA, a single institution study is inherently limited by a smaller number of patients that can be accrued compared to a multi-institutional study design. This limitation is magnified with the relative rarity of ORN. A second important consideration is the duration of or follow up period. Given that ORN is a late complication of EBRT, there are likely additional patients in our cohort that may go on to develop this disease in the future. This has possibly skewed our analysis of the interval of time from the initiation of EBRT to diagnosis of ORN as those with longer intervals would not be captured by our study. We believe that the most accurate risk modelling for ORN development will need to account for disease site (oral cavity and oropharynx with greatest ORN risk) and stage, overall dental status and health, radiation dose and also time from radiation in order to counsel patients in the most effective manner.

## Data Availability

All data produced in the present work are contained in the manuscript.

## Notes

Funding source: This material is the result of work supported with resources and the use of facilities of the Michael E. DeBakey VA Medical Center. VCS is supported by a Career Development Award from the Veterans Administration Clinical Science Research and Development division (1IK2CX001953).

Conflicts of interest: Contents do not represent the views of the US Department of Veterans Affairs or the US Government. The authors report no conflicts of interest for the existing work.

### Competing Interest Statement

The authors have declared no competing interest.

### Funding Statement

This material is the result of work supported with resources and the use of facilities of the Michael E. DeBakey VA Medical Center. VCS is supported by a Career Development Award from the Veterans Administration Clinical Science Research and Development division (1IK2CX001953).

### Author Declarations

Following approval from Baylor College of Medicine and Michael E. DeBakey Veterans Affairs Medical Center (MEDVAMC) Institutional Review Boards, we reviewed the records of patients who were diagnosed and initiated radiation therapy at the MEDVAMC between January 2016 and December 2020 captured in our treatment tracking registry.

